# Infection Dynamics of Coronavirus Disease 2019 (Covid-19) Modeled with the Integration of the Eyring’s Rate Process Theory and Free Volume Concept

**DOI:** 10.1101/2020.02.26.20028571

**Authors:** Tian Hao

## Abstract

The Eyring’s rate process theory and free volume concept, two very popular theories in chemistry and physics fields, are employed to treat infectious disease transmissions. The susceptible individuals are assumed to move stochastically from one place to another. The virus particle transmission rate is assumed to obey the Eyring’s rate process theory and also controlled by how much free volume available in a system. The transmission process is considered to be a sequential chemical reaction, and the concentrations or fractions of four epidemiological compartments, the susceptible, the exposed, the infected, and the removed, can be derived and calculated. The obtained equations show that the basic reproduction number, *R*_0_, is not a constant, dependent on the volume fraction of virus particles, virus particle size, and virus particle packing structure, the energy barrier associated with susceptible individuals, and environment temperature. The developed models are applied to treat coronavirus disease 2019 (Covid-19) transmission and make predictions on peak time, peak infected, and *R*_0_. Our work provides a simple and straightforward approach to estimate how infection diseases evolve and how many people may be infected.

## I. INTRODUCTION

Infectious diseases occur frequently within last twenty years. The 2003 SARS (*Severe Acute Respiratory Syndrome*), 2009 influenza A (*H*1*N*1), 2013 ebola virus, and current coronavirus inflected pneumonia (*Covid* − 19) are among the deadliest and have posed a strong impact on human lives and activities. Mathematical modeling and analysis of infectious disease transmissions^1–7^ have been performed for a century. Its importance in understanding infectious disease transmissions and developing practical strategies for disease control and mitigation have been demonstrated. Challenges remain in difficult descriptions of human contact patterns and virus evolution, due to randomness of human interactions and unpredictability of virus growth especially for a new type of virus. The problems encountered in disease transmission area are similar to many-body problems encountered in chemistry and physics fields, where many entities like electrons, atoms, molecules, or nano-sized even micro-sized particles interact each other in a disordered manner.

The Eyring’ rate process theory^8^ and free volume concept^9–13^ have been employed to solve many-body problems without involving complicated quantum mechanical calculations. The Eyring’s rate process theory, originated from quantum mechanics, argues that every physical or chemical phenomenon is a rate controlled process, such as chemical reactions, electron transfer, conductivity, viscous flows, diffusions, etc. Every process needs an activated energy to move from the initial state to the final state. Free volume theory, originated from molecular systems, is a most successful mean field theory in dealing with many-body problems. All different kinds of interactions among entities are factored into a single term, the free volume available in a system. Since the free volume theory resolves how large freedom an entity may have and the Eyring’s rate process theory describes how fast the process is, these two theories have been integrated together to describe many seemingly unrelated systems or phenomena like glass liquids^12^, colloids and polymers^14,15^, granules^16–18^, electrical and proton conductivity^19,20^, superconductivity^21^, and Hall Effect^22^ with great success.

In this article, we will utilize the Eyring’s rate process theory and free volume concept to describe infectious disease transmission process. The infectious disease transmission is considered as a sequential chemical reaction. We will follow the popular SIR (*susceptible, infectious, and removed*) and SEIR (*susceptible, exposed, infectious, and removed*) compartment categorization methods proposed in the literature^1–7^ to develop our theory. The difference between these two models can be found in literature^4^. SEIR model can give more information like latency period. Each category will be treated as a “chemical reactant”. Its concentration or fraction expressed in the number of individuals in each segment divided by the total population in consideration will be estimated for unveiling disease transmission mechanisms and predicting peak time and peak infected population.

## II. THEORY

Considering that some of the recovered individuals may be infected again, we therefore add a backward reaction in the last step of the sequential chemical reaction. The whole infection process can be expressed as below:

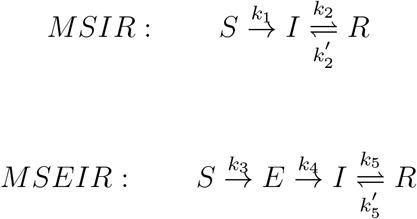

The only difference between SIR and SEIR models is the initial step: SIR model assumes that the susceptible will directly transform into the infected, while SEIR model assumes that there is an intermediate state “exposed”. For avoiding confusion with traditional SIR and SEIR models, we will call our modified versions as MSIR and MSEIR in this article. The key is how to theoretically determine chemical reaction rate constants like 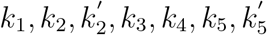 expressed in two chemical reactions shown above. Once these parameters are known, the fractions of *S, E, I, R* can be predicted. The first step in MSIR model and the second and third steps in MSEIR model are one way irreversible chemical reactions. The last step in both MSIR and MSEIR models are reversible. Note that both 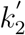 and 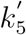 should be very small and the initial fraction of the recovered should be small too, leading to a negligible contribution to the infected pool.

For the first step reaction, assume that susceptible individuals move randomly and become “exposed” first. The virus particles then transmit from one individual to another. This individual thus becomes “infected”. So in MSIR model, there should be two steps involved during the process from “*S*” to “*I*”: the individual movement and virus particle movement. Human movement is a stochastic random process and should be similar to the movements of atoms, molecules, colloidal particles, and even granular powders, which have been studied extensively. Someone may argue that human movement is an athermal process, while atomic, molecular, and colloidal particle movements are thermal driven. For athermal granular powder under a tapping process, we theoretically found that they behave like thermal systems and follow the stretched exponential pattern in term of tap density changing with the number of taps^16^. Our approach is mainly based on Theodor F*ö*rster’s theory^23,24^ that deals with the energy transfer from donors to random distributed acceptors. Theodor F*ö*rster’s theory involves a very complicated quantum mechanical treatment but its mathematical backbone was simplified and extracted by Blumen^25^ and Klasfter^26^. Detailed information can be found in my previous article^16^. Furthermore, human collective motion and individual walking behaviors of animals were found to behave like thermal systems and follow Boltzmann distribution, though the temperature needs to be defined differently in these athermal systems^27,28^. Utilizing the same approach to treat “human movements” in disease transmissions, we may write the reaction constant as^16^:

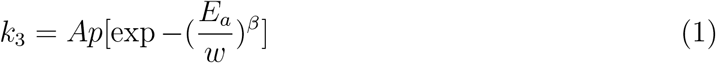

where *A* and *p* are constant, *β* is stretched exponential parameter of values between 0 and 1, *E*_*a*_ is the energy barrier for a human to move around, and *w* is the basic energy that a person may need during a normal circumstance.

After an individual is exposed, the transmission of virus particles from one person to another will make an “exposed” person become “infected”. This process will be dependent on the free volume available for virus particles to travel and how fast these virus particles will travel. It should be similar to the viscosity or conductivity of an entity that has been addressed in many systems in my previous articles^12,14,19,20,29^. The free volume can be estimated with inter-particle spacing concept^14,30^. For infectious diseases, more free volume for both human and virus particles means “isolation” and less transmission. High virus particle volume fractions mean higher transmissions, which is similar to viscosity of colloidal suspensions or granular powders. Therefore the chemical reaction rate should be proportional to the “viscosity” of this entity, then *k*_4_ can be written as^29^:

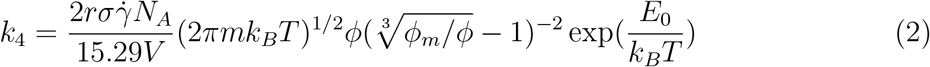

where *V* is the volume in consideration, *r* is the radius of virus particles, *σ* is the shear stress applied when virus particles transmit from one place to another, 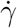 is the shear rate, *N*_*A*_ is Avogadro number, *m* is the mass of a virus particle, *k*_*B*_ is Boltzmann constant, *T* is the temperature, *ϕ* is the volume fraction of virus particles in the volume *V, ϕ*_*m*_ is the maximum packing fraction of virus particles, and *E*_0_ is the energy barrier for virus particles.

In MSIR model, we assume that during the transmission process from the susceptible to the infected, both human movement and virus particle transmission are involved and the “exposed” is only an transient state. According to the transient state theory of chemical reaction^8^, we may easily obtain:

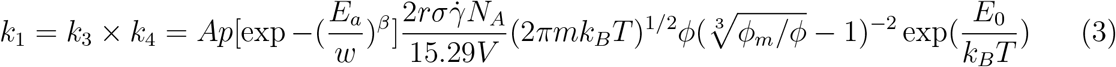

Eq.(1-3) indicate that infectious disease transmission is a complicated process, and is dependent on many factors like human movement energy barrier, the particle size and volume fraction of virus particles, the mass of a virus particle, temperature, and volume in consideration. Smaller volume leads to lower transmission constant, and isolation definitely is a good method to preventing virus from spreading.

For a sequential chemical reaction, the fraction of each reactant can be expressed with a series of differential equations^31^. For MSIR model, we may write:

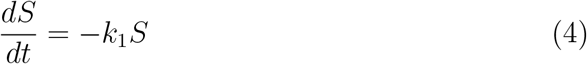

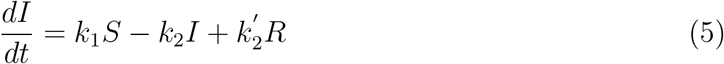

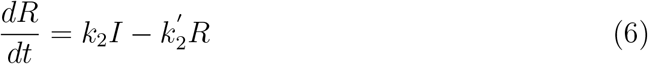

For MSEIR model, we may write:

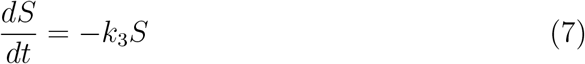

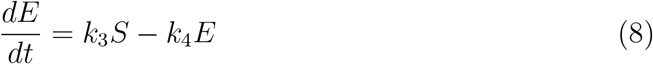

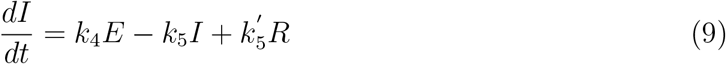

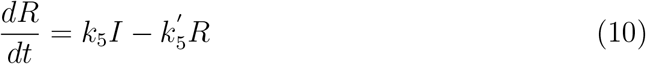

Assume that the initial fraction of the susceptible is *S*_0_, we may easily obtain:

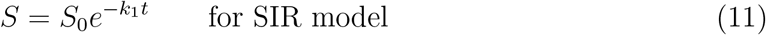

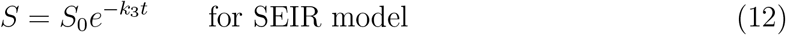

Since the contribution from 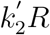 to the infected is negligible based on the fact that the recovered may gain immunity from the disease and the fraction of the recovered is relatively small at early stages, the first step reaction product, *E* and *I* may be written:

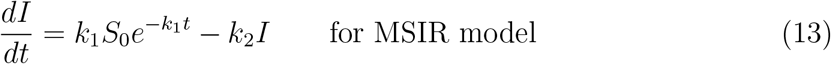

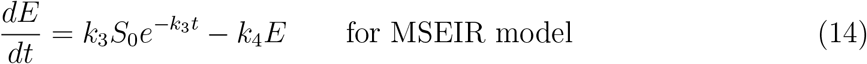

Both equations above are first-order differential equations of standard form:

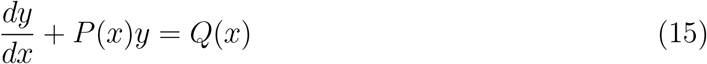

which has a standard solution as:

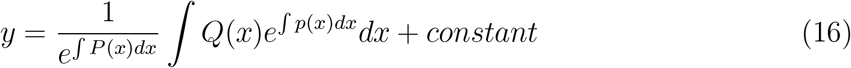

We therefore obtain:

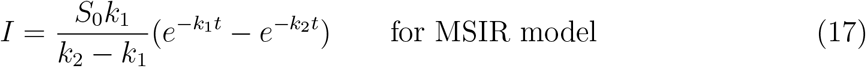

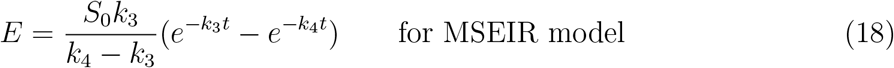

Similarly, ignoring the contribution from 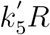, we may obtain *I* in MSEIR as shown below:

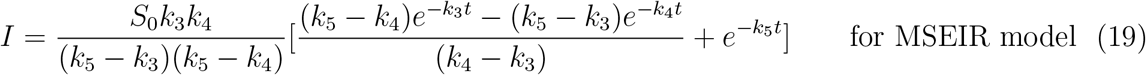

Assuming that *S* + *I* + *R* = *S*_0_ for MSIR model and *S* + *E* + *I* + *R* = *S*_0_ for MSEIR model, we can obtain:

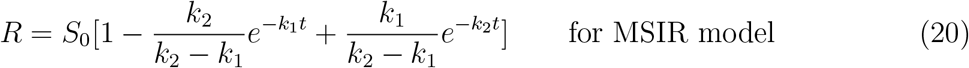

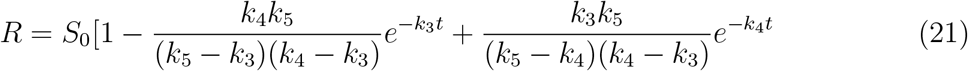

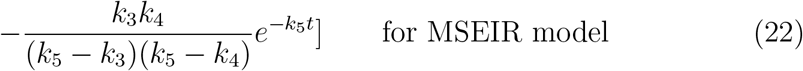

Both *I* and *E* should have a peak value that can be simply determined by differentiating Eq. 18 against time:

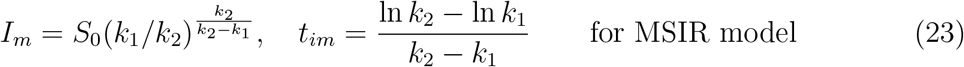

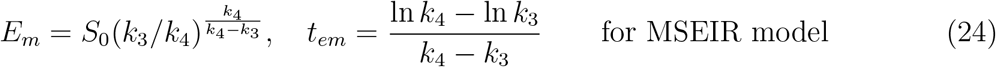

In theory, the peak value of *I* in MSEIR model can be determined with a similar way but there is no exact analytical solution. We may determine approximate peak values after the equations are plotted out.

A very important parameter in infectious disease transmission is the basic reproduction number, *R*_0_, which is the transmission rate divided by the recovery rate. It is considered to be an indicator on the severity of infectious transmissions. Note that our approach is different from original Kermack-McKendrick SIR model^1,4^ and SEIR model^4,32,33^. In these models,

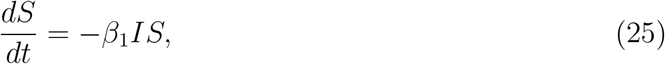

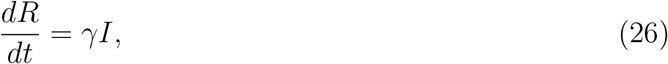

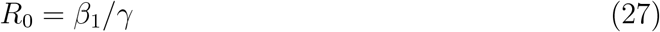

if the death rate is not considered. Comparing these equations with our models expressed in Eq. 6 and Eq. 10, we may easily obtain *R*_0_ as below:

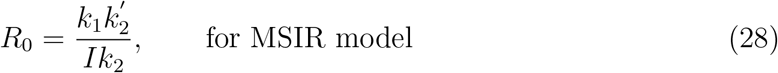

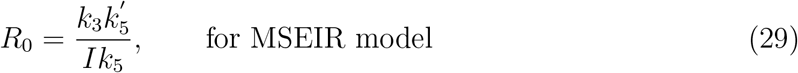

For a reversible chemical reaction like the last step reaction between the infected and the removed, an equilibrium should be reached and the chemical reaction rate constant is 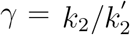 for MSIR model and 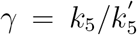 for MSEIR model. Clearly, *R*_0_ actually is not a constant, dependent on temperature and virus particle volume fraction. So far we already know *k*_1_, *k*_3_, *k*_4_. The rate constants like 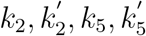 can be determined with the recovered data that are relatively easy to be counted during the initial period of virus outbreak. We then can predict the peak time, the peak infected, and the basic reproduction number, etc.

## III. RESULTS

The fractions of the susceptible, exposed, infected, and recovered can be computed as a function of time based on the equations above and shown in Figure 1 for both MSIR and MSEIR models. The exposed and the infected peak at certain time and the recovered increases during the same time of period, which is consistent with observations and other models in the literature.

**Figure 1:**
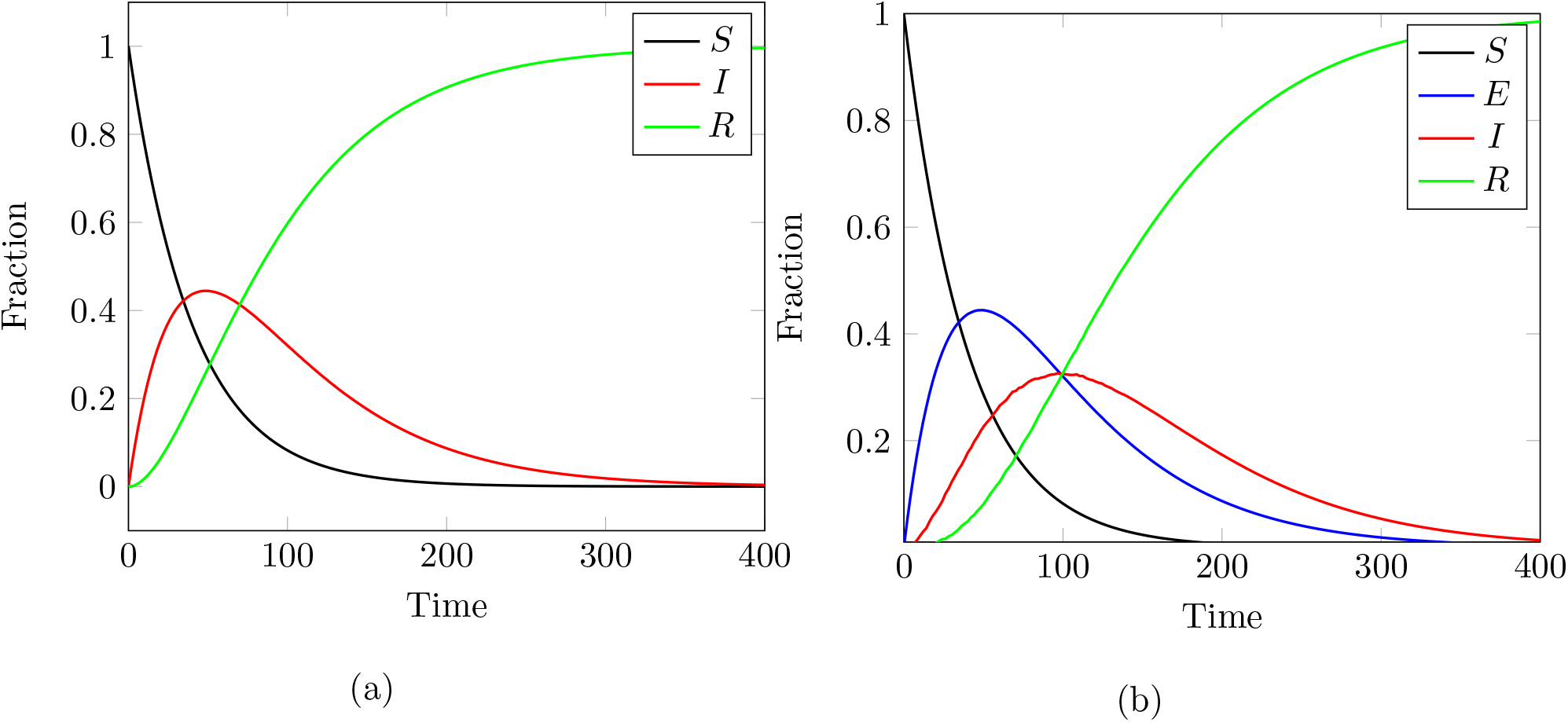
Fraction of each categorical individuals against time in (a) MSIR model with 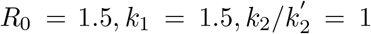;(b) MSEIR model with 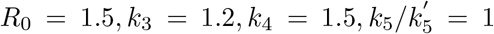. We intentionally assign similar parameters for these two models for comparing epidemic difference. MSIR model predicts a earlier and higher infectious peak position than MSEIR model. *S*_0_ is assumed to be 1.

Since *k*_1_ and *k*_4_ are functions of virus particle volume fraction and environment temperature, the fraction in each category should change with these two parameters, too. Besides virus particle volume fraction and temperature related terms in Eq. 2 and Eq. 3, the remaining is related to virus particle radius and mass, and the spacial volume under consideration. Clearly, large virus particle size and mass lead to large rate constants. For clearly showing the relationship between rate constants and virus particle volume fraction and temperature, we may rearrange Eq. 2 and Eq. 3 as below:

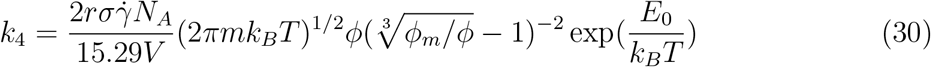

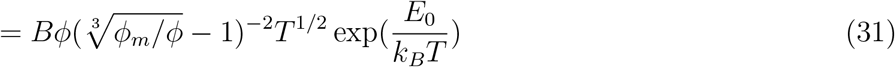

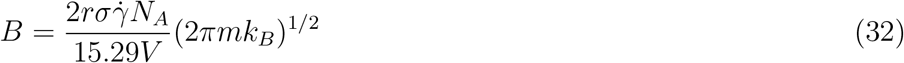

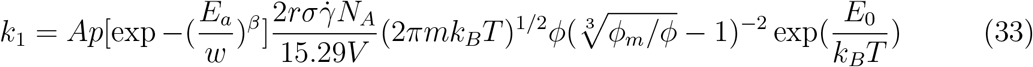

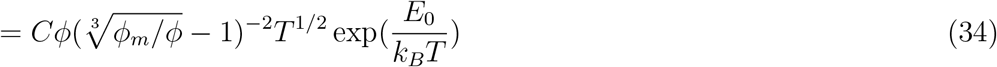

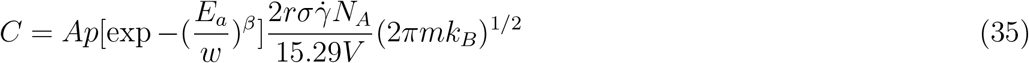

*B* and *C* are a constant for a specific virus and a spacial volume, when virus particles are under normal shear forces during a transmission. For simplification reason, we may assign a numeric value to both *B* and *C*, thus the impact of virus particle volume fraction and temperature on the fraction of the infected can be evaluated illustratively. The infected against virus particle volume fraction is plotted in Figure 2. A much stronger dependence of virus particle volume fraction is predicted with MSIR model than MSEIR, possibly due to the buffering effect caused by the “exposed” category in the latter. For MSIR model, an explosive increase of the infection is predicted when virus particle volume reaches about 45%. It looks like a percolation transition phenomenon^34,35^, a first-order phase transition frequently observed in amorphous multi-phase systems like colloidal suspensions, where contacts and connections between individual entities play a critical role in controlling physical properties of whole system. The relationship between the infected and virus particle volume fraction seems to be reasonable, as intuitively more infection is expected if many virus particles are present in a system. Some researchers have already claimed that infectious disease transmissions are a percolation phenomenon in nature^36^ and applied percolation theory to study epidemic spreading^36,37^. Our prediction is in line with those research work.

**Figure 2:**
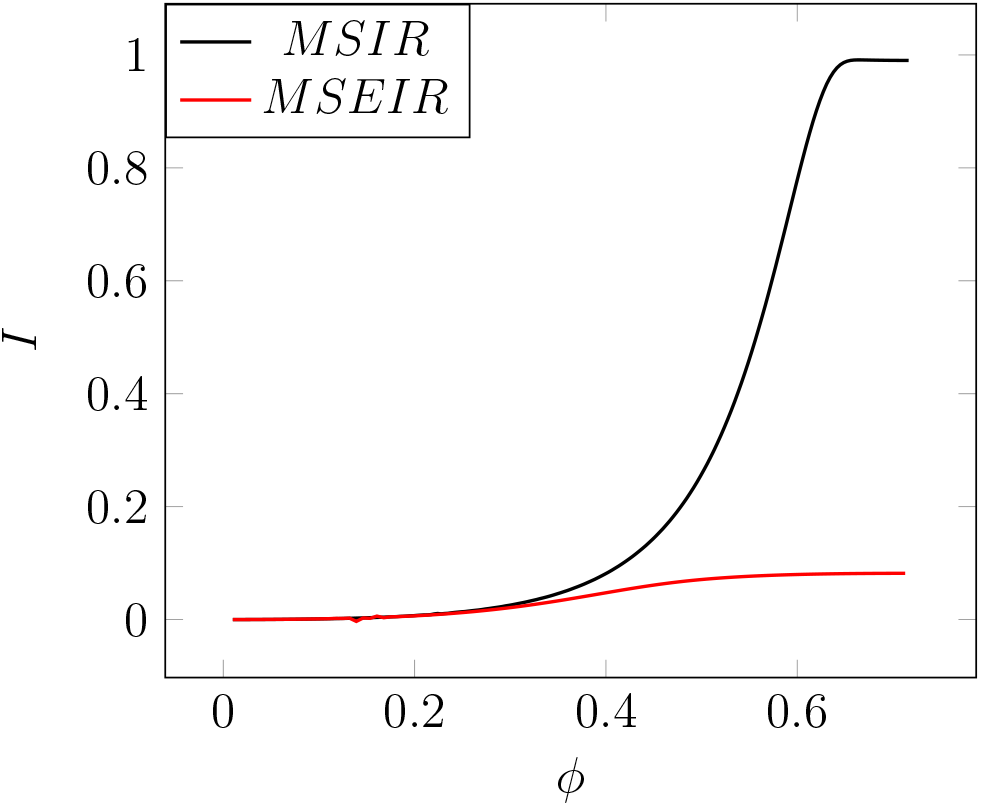
Fraction of the infected against virus particle volume fraction in (a) MSIR model with 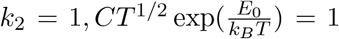 (b) MSEIR model with *k*_3_ = 1.5, *k*_5_ = 1, *ϕ*_*m*_ = 0.72, the maximum volume packing fraction of face-centered cubic packing structure, 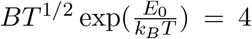. MSIR model predicts a stronger dependence on virus particle volume fraction than MSEIR model. *S*_0_ is assumed to be 1, too.

The infected is not only dependent on virus particle volume fraction, but also dependent on the temperature, which is shown in Figure 3. The energy barrier term *E*_0_ of course determines how temperature may impact disease transmission rate thus the number of the infected. The infected in MSEIR model shows a much stronger temperature dependence and decreases rapidly with temperature changing from 0 to 60 °C, while the infected in MSIR model just decreases a little bit with temperature. Higher temperatures definitely can reduce disease transmission. From past observations, infectious disease transmissions typically show a seasonal sinusoidal variation. Most infectious diseases like influenza happen in winter time and diminish when spring or summer comes in. Coronavirus (Covid-19) pandemics initially started some time in December 2019 at Wuhan, China, has infected more than 75000 people with over 2200 deaths so far. How long time this infectious transmission is going to last and when it will peak are critical information to make proper mitigation strategy. Utilizing the data provided by Chinese Center for Disease Control and Prevention (CCDC) from Jan. 21 to Feb.17, 2020, the fraction of the infected is predicted with both MSIR and MSEIR models and shown in Figure 4. From these data, we are able to determine the reaction rates at each step and make predictions. As shown in Fig.4 (b), both models predict that the infected may peak about 150 days from January 21, 2019. For MSIR model, the exact peak time and the peak values can be calculated with Eq. 24 with the parameters obtained from Fig.4, *t*_*m*_ = 158 days and *I*_*m*_ = 0.854, corresponding to 170826 infected individuals. For MSEIR model, we cannot have exact solutions to the equation but can estimate using Fig. 4 (b): the peak time is a little longer than the predicted with MSIR model, about 160 days, and has a higher infected fraction about 0.9, corresponding to about 180000 infected individuals. Both models predict a skewed distribution, implying that the fraction of the infected remains at high ends for a relative long time. Please keep in mind that these predictions should be strongly dependent on how the isolation measures are executed and how accurate the data are, which may affect the rate constants used in the calculations.

**Figure 3:**
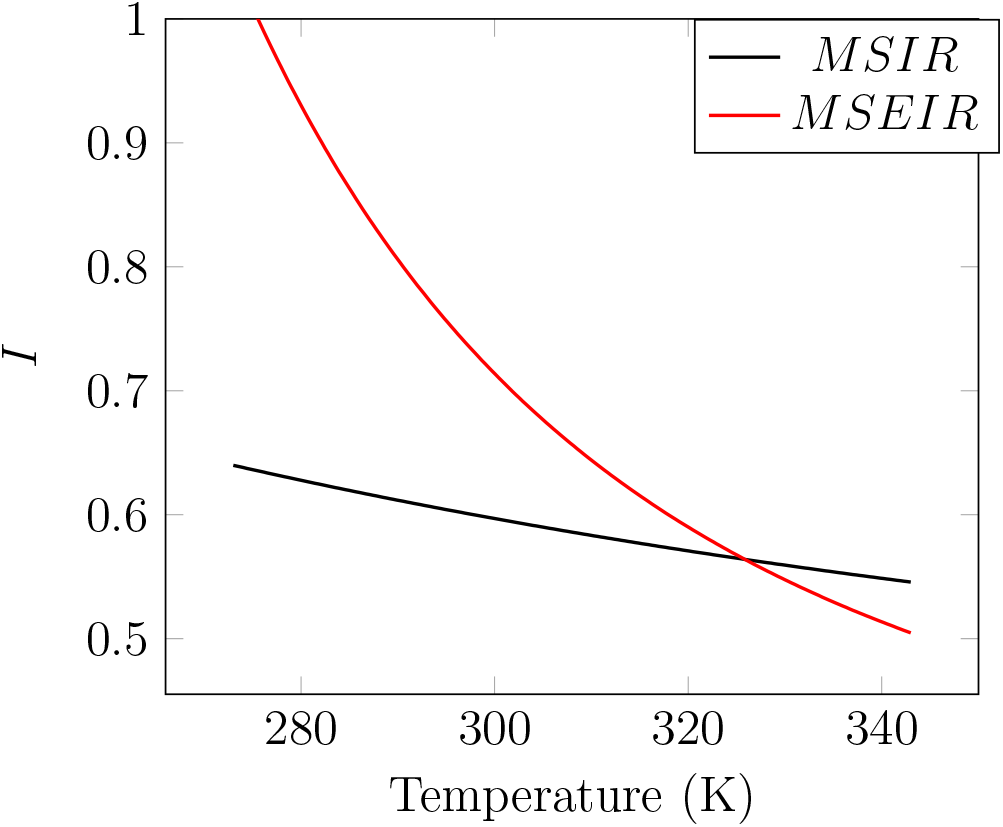
Fraction of the infected against temperature in (a) MSIR model with *k*_2_ = 1;(b) MSEIR model with *k*_3_ = 1.5, *k*_5_ = 1. MSEIR model predicts a stronger temperature dependence than SIR model. For simplification reason, the other terms irrelevant to temperature in Eq. 32 and Eq. 35 are assumed as 1. *S*_0_ is assumed as 1, too.

**Figure 4:**
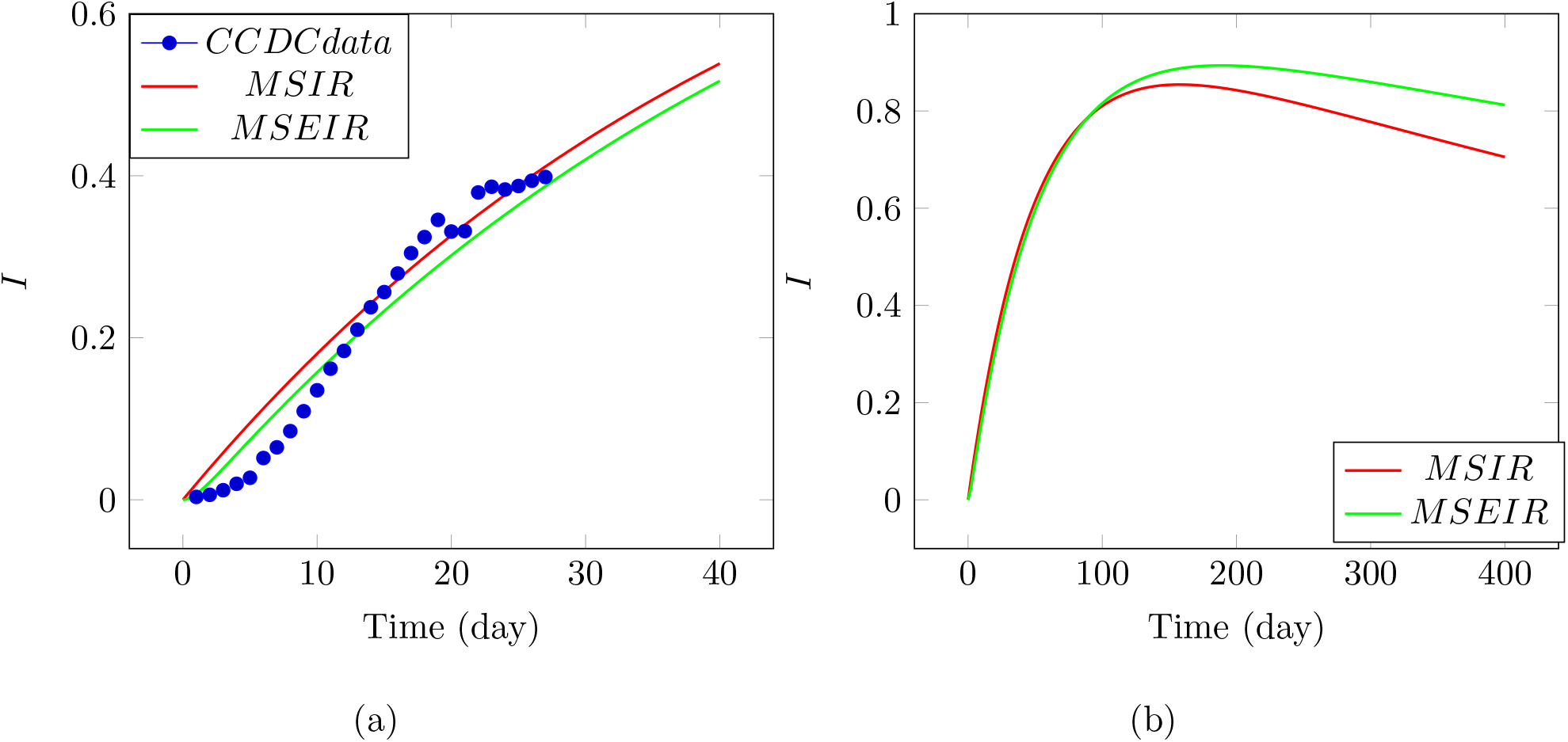
The fraction of the infected based on coronavirus disease (Covid-19) data provided by Chinese Center for Disease Control and Prevention (CCDC) from Jan. 21 to Feb.17, 2020, and regressed with MSIR and MSEIR models. The number of the infected individuals is divided by a population size 200000 to obtain the fraction. (a) The real data points and predicted with both MSIR and MSEIR models. (b) The same equations but plotted in a wider range. For MSIR model, *k*_1_ = 0.001, *k*_2_ = 0.02. For MSEIR model, *k*_3_ = 0.019, *k*_4_ = 1.05, *k*_5_ = 6 *×* 10^−4^. *S*_0_ = 1 is assumed as 1 in both cases.

The fraction of the recovered is important to know, as it directly relates to how many people may survive from this epidemics. The original SIR and SEIR models only consider “the removed” that may contain both the recovered and the death. Figure 5 shows the recovered (a) and the recovered plus the death (b) against time, predicted with both MSIR and MSEIR models. Theoretically, the same rate constants obtained from the infected data in Fig. 4 should be used for making predictions for the recovered in Fig. 5. Unfortunately, these two groups of data provided by CCDC don’t match each other and cannot be integrated into one model. If the same parameters like *k*_1_, *k*_2_, *k*_3_, *k*_4_, *k*_5_ obtained from the infected data were used for the removed data regressions, the predicted with MSIR and MSEIR models were 8 and 13 times smaller than the reported, respectively, no matter that the recovered only or the addition of the recovered and the death is used. In average, there is almost 10 times difference between these two group of data. If the removed data are accurate, then the infected should be 10 times larger than the reported, which turns out that 724360 people should be infected by Feb. 17, 2020. Note that the regressions are relatively poor for the recovered and even worse for the recovered plus the death. The long latency period and lack of an effective drug to cure the disease may add complexity of claiming a patient to be fully recovered for this new kind of disease, which could contribute to the poor regression.

**Figure 5:**
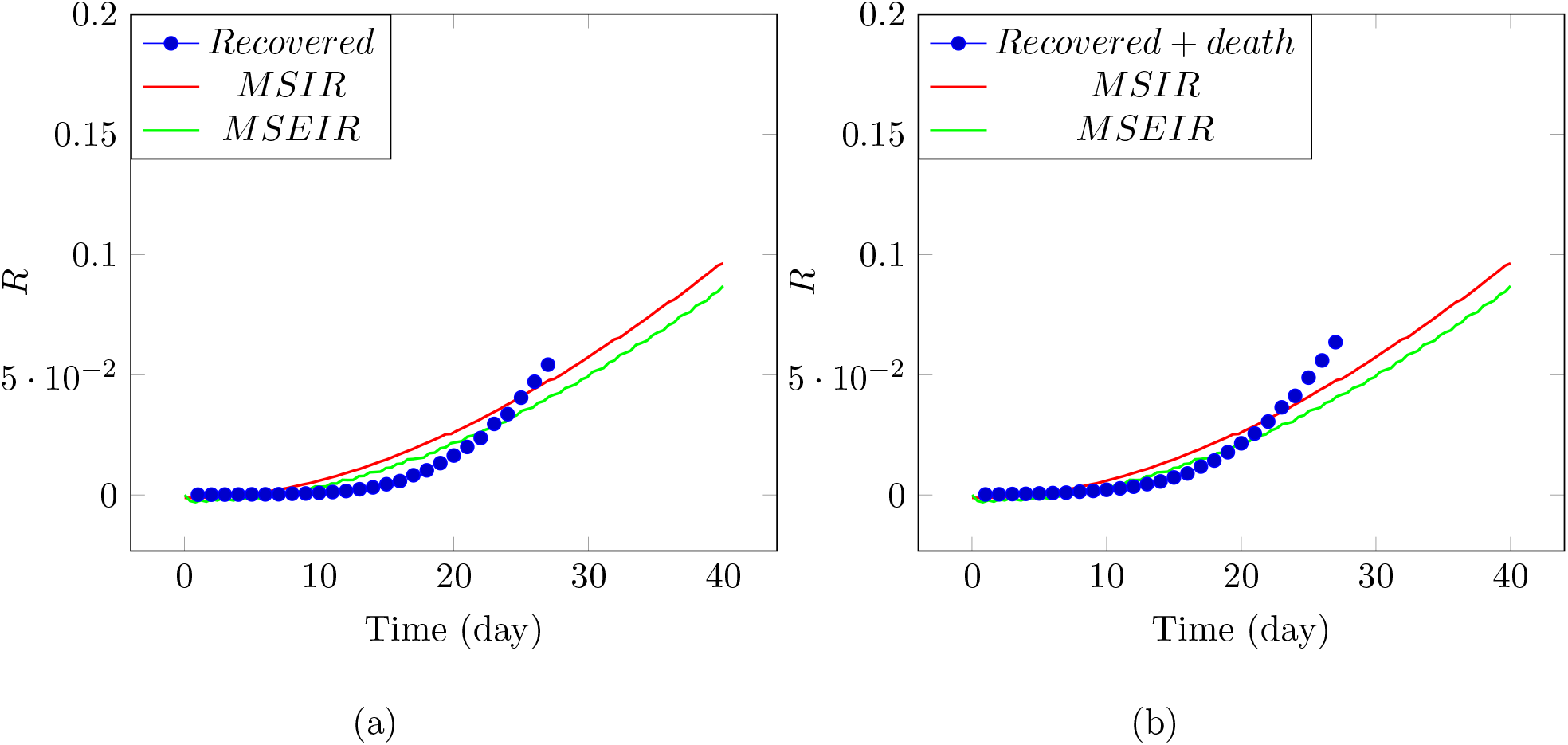
The fraction of the removed based on coronavirus disease (Covid-19) data provided by Chinese Center for Disease Control and Prevention (CCDC) from Jan. 21 to Feb.17, 2020, and regressed with MSIR and MSEIR models. The population size 200000 was used for calculating the fractions. (a) Only the recovered data are used for modeling; (b) Both the recovered and the death data are added together for modeling. Same parameters obtained from the infected data are used for calculations. For MSIR model, *k*_1_ = 0.02, *k*_2_ = 0.001, *S*_0_ is assumed to be 1. For MSEIR model, *k*_3_ = 0.019, *k*_4_ = 1.05, *k*_5_ = 0.0006, *S*_0_ is assumed to be 1.

Another critical parameter in infectious disease is the basic reproduction number, *R*_0_, which can be computed with Eq. 29. With the parameters obtained from the infected data provided by CCDC, the calculated *R*_0_ is plotted against time in Figure 6. Initially, *R*_0_ is really high, but decays rapidly and approaches a steady value. The predicted steady *R*_0_ value is about 3.5 according to MSEIR model and about 1.5 according to MSIR model. Early studies on coronavirus show *R*_0_=2.2^38^, 2.68^40^, and 3.11^39^. Variation of *R*_0_ may stem from the fact that it is not a constant in theory based on our model. Complex estimation methods on *R*_0_ make some researchers believe that *R*_0_ is not an indicative parameter to use^4,32,33^. However, our method is straightforward and can potentially be used to standardize *R*_0_ calculation.

**Figure 6:**
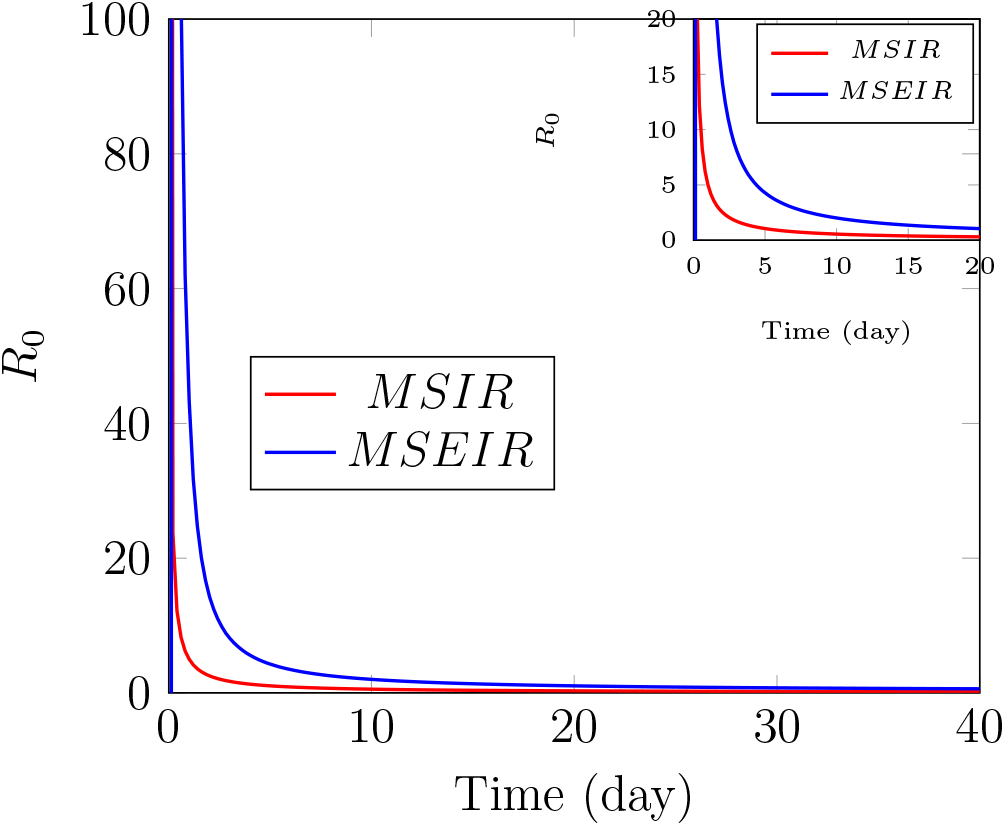
*R*_0_ of coronavirus Covid-19 calculated with MSIR and MSEIR models. The parameters are same as used in the infected regression in Fig.4. For MSIR model, 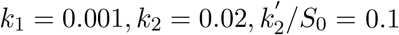. For MSEIR model, 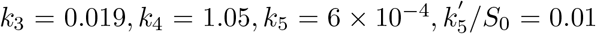. The inlet is the same graph but plotted in a smaller scale.

In summary, using same categorical methods as in traditional SIR and SEIR models, we consider infectious disease transmissions to be a sequential chemical reaction and treat the transmission process with the integration of both Eyring’s rate process theory and free volume concept together. Based on the data provided by CCDC, the peak time, the peak infected, the total infected based on the recovered data, and *R*_0_ are estimated and shown in table I. These data indicate that current epidemics may last more than a year and total infected people may be between 579488 and 941668.

**Table I:**
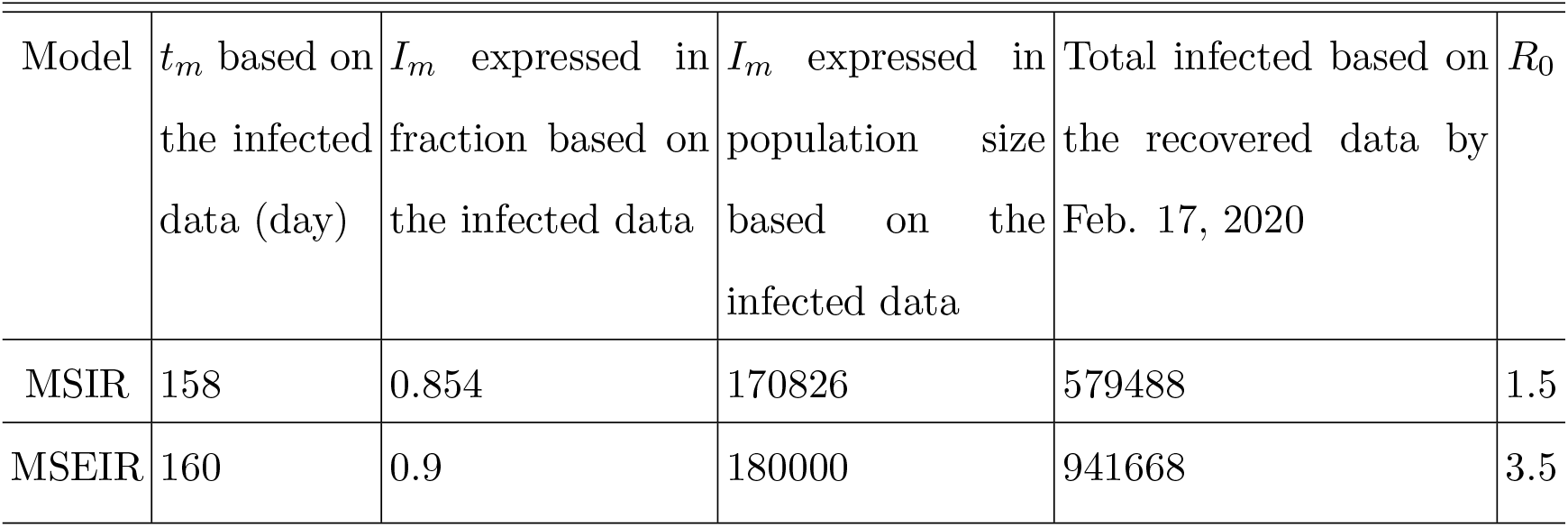
Peak time, peak infected, *R*_0_ estimated with MSIR and MSEIR models

| Model | $t_m$ based on the infected data (day) | $I_m$ expressed in fraction based on the infected data | $I_m$ expressed in population size based on the infected data | Total infected based on the recovered data by Feb. 17, 2020 | $R_0$ |
| --- | --- | --- | --- | --- | --- |
| MSIR | 158 | 0.854 | 170826 | 579488 | 1.5 |
| MSEIR | 160 | 0.9 | 180000 | 941668 | 3.5 |

## IV. DISCUSSION

Disease transmission is difficult to describe, as it is hard to identify the sources of diseases, confirm infected individuals, predict human movement pattern and virus particles transmission pattern, etc. The physical mechanisms and principles governing infectious disease transmissions should be similar to the observed in amorphous disorder systems, such as the percolation transition, Boltzmann distribution probability, the stretched exponential relaxation, etc. Human motion is not a thermal driven movement, however, has been found to follow the same Boltzmann distribution probability. The volume fraction of virus particles should lead to a dramatic increase in transmission rate, similar to percolation transition process. The common many-body problems encountered in chemistry and physics world can be resolved with the integration of Eyring’s rate process theory and free volume concept, which has been demonstrated to work well for many multi-scale systems ranging from electrons to granular particles, even the universe^41^. This approach is therefore naturally applied to the disease transmissions. Attempt made in this article is to understand physical mechanisms of infectious disease transmissions using same categorical methods popularly employed in infectious disease transmission field. Although our focus is put on coronavirus (Covid-19) currently spreading around the world, the theoretical framework proposed in this article can be applied to other transmission diseases, too.

The successful predictions are dependent on data accuracy. For every new type of a virus, the test, identification, diagnostic, and counting methods need time and effort to be properly established. It is fully understandable that the initial data don’t reflect the true epidemic process. We utilize the official data released by CCDC to test our models. The predicted parameters shown in Table I should be used cautiously.

Our approach is relatively simple and straightforward without involving complicated mathematical calculations, in comparison with other theoretical models available in the literature. Extra seasonal force doesn’t need to be inserted into our model for demonstrating the seasonality of infectious diseases, as rate constants are temperature dependent.

## V. CONCLUSION

A sequential chemical reaction process from the susceptible, to the exposed, the infected, and the removed in the end is proposed to describe disease transmissions. At each step, there is a chemical rate constant associated with and the fraction of each product/reactant can be easily calculated. Every step is a one-way reaction, except the last step from the infected to the removed, as some small amount of the removed could be infected again. The Eyring’s rate process theory and free volume concept continue to be used for defining rate constants.

In general, our model predicts that the fraction of the infected will peak at certain time, increase with the volume fraction of virus particles, and decrease with temperature. The energy barrier for human to move has played a critical role in disease transmissions. The isolation process with a huge energy barrier for human can change the chemical reaction rate constant dramatically. The basic reproduction number is not a constant and can be computed once the parameters in the models are determined.

In particular, we applied our models to treat coronavirus (Covid-19) transmission. Based on the data provided by CCDC, we predict that the infected may peak at 158 days from Jan. 21, 2020 and may take a long time, more than a year, to drop to zero, though MSIR and MSEIR models give a slightly different estimation. The data of the infected don’t match with the removed for both models: based on the infected data, the removed is predicted to be 8 and 13 times smaller than the actual reported according to MSIR and MSEIR models. If the removed data, either the recovered or the recovered plus the death, are reliable, the actual infected people is between 579488 and 941668 by Feb. 17, 2020. The basic reproduction number of coronavirus (Covid-19) is computed to be about 1.5 based on MSIR model and 3.5 based on MSEIR model.

## Data Availability

Data are provided by CCDC

## Acknowledgments

The author sincerely appreciate colleagues’ and reviewers’ feedback and comments for substantially improving the readability and rationality of this article.

## References

1. W. O. Kermack and A. G. McKendrick, A contribution to the mathematical theory of epidemics, Proc. R. Soc. Lond. A, 1927, 115, 700–721

2. J. L. Aron, Seasonality and Period-doubling Bifurcations in an Epidemic Model, J. Theor. Biol., 1984, 110, 665–679

3. N. C. Grassly and C. Fraser, Mathematical models of infectious disease transmission, Nature Reviews Microbiology, 2008, 6, 477–487

4. B. Ridenhour, J. M. Kowalik, and D. K. Shay, Unraveling R_0_: considerations for public health applications, Am J Public Health, 2014, 104(2), 32–41

5. Cesar Parra-Rojas, Thomas House, and Alan J. McKane, Stochastic epidemic dynamics on extremely heterogeneous networks, Phys. Rev. E, 2016, 94, 062408

6. C. I. Siettos and L. Russo, Mathematical modeling of infectious disease dynamics, Virulence, 2013, 4(4), 295–306

7. Alun L. Lloyd, Realistic Distributions of Infectious Periods in Epidemic Models: Changing Patterns of Persistence and Dynamics, Theoretical Population Biology, 2001, 60, 59–71

8. S. Glasstone, K. Laidler, and H. Eyring, The Theory of Rate Process, McGraw-Hill, 1941

9. M. H. Cohen and D. Turnbull, Molecular transport in liquids and glasses, J. Chem. Phys., 1959, 31, 1164–1169

10. D. Turnbull and M. H. Cohen, Free-Volume Model of the Amorphous Phase: Glass Transition, J. Chem. Phys., 1961, 34, 120–124

11. J. C. Dyre, Source of non-Arrhenius Average Relaxation Time in Glass-forming Liquids, J. Non-Crystalline Solids, 1998, 235-237, 142–149

12. T. Hao, Unveiling the Relationships among the Viscosity Equations of Glass Liquids and Colloidal Suspensions for Obtaining Universal Equations with the Generic Free Volume Concept, Phys. Chem. Chem. Phys., 2015, 17, 21885–21893

13. H. Fujita, Notes on Free Volume Theories, Polymer J., 1991, 23, 1499–1506

14. T. Hao, Viscosities of liquids, colloidal suspensions, and polymeric systems under zero or non-zero electric field, Adv. Coll. Interf. Sci., 2008, 142, 1–19

15. T. Hao, Electrorheological Fluids: The Non-aqueous Suspensions, Elsevier, December 19, 2005

16. T. Hao, Derivation of stretched exponential tap density equations of granular powders, Soft Matter, 2015, 15, 3056–3061

17. T. Hao, Tap density equations of granular powders based on the rate process theory and the free volume concept, Soft Matter, 2015, 11, 1554–1561

18. T. Hao, Defining Temperatures of Granular Powders Analogously with Thermodynamics to Understand the Jamming Phenomena, AIMS Materials Science, 2018, 5(1), 1–33

19. T. Hao, Electrical Conductivity Equations Derived with the Rate Process Theory and Free Volume Concept, RSC Adv., 2015, 5, 48133–48146

20. T. Hao, Conductivity Equations of Protons Transporting Through 2D Crystals Obtained with the Rate Process Theory and Free Volume Concept, Chem. Phys. Lett, 2018, 698, 67–71

21. T. Hao, Exploring high temperature superconductivity mechanism from the conductivity equation obtained with the rate process theory and free volume concept, Chem. Phys. Lett, 2019, 714, 99–102

22. T. Hao, Integer, Fractional, and Anomalous Quantum Hall Effect Explained with Eyring’s Rate Process Theory and Free Volume Concept, Phys. Chem. Chem. Phys, 2017, 19, 6042–6050

23. T. Förster, Energiewanderung und Fluoreszenz, Naturwissenschaften, 1946, 33, 166 – 175; English translation, T. Förster, Energy migration and fluorescence, J. Biomed. Opt., 2012, 17, 011002

24. T. Förster, Expermentelle und theoretische Untersuchung des zwischengmolekularen Ubergangs von Elektronenanregungsenergie, A Naturforsch, 1949, 4A, 321–327

25. A. Blumen, Excitation transfer from a donor to acceptors in condensed media: a unified approach, Il Nuovo Cimento, 1981, 63, 50–58.

26. J. Klafter and M. F. Shlesinger, On the relationship among three theories of relaxation in disordered systems, Proc. Natl. Acad. Sci. U. S. A.,, 1986, 83, 848–851.

27. J. L. Silverberg, M. Bierbaum, James P. Sethna, Itai Cohen, Collective Motion of Humans in Mosh and Circle Pits at Heavy Metal Concerts, Phys. Rev. Lett., 2013, 110, 228701

28. S. Petrovski, A. Mashanova, and V. A. A. Jansen, Variation in individual walking behavior creates the impression of a Lévy flight, Proc Natl Acad Sci U S A., 2011, 108(21), 8704–8707.

29. T. Hao, Analogous Viscosity Equations of Granular Powders Based on Eyring’s Rate Process Theory and Free Volume Concept, RSC Adv., 2015, 5, 95318–95333

30. T. Hao, and R. E. Riman, Calculation of Interparticle Spacing in Colloidal Systems, J. Coll.Interf. Sci., 2006, 297, 374–377

31. P. Atkins and J. de Paula, Physical Chemistry, 8th Edition, WH Freman, 2006

32. P. den Driessch, Reproduction numbers of infectious disease models, Infectious Disease Modelling, 2017, 2, 288–303

33. J. Li, D. Blakeley, and R. J. Smith, The Failure of R_0_, Comput. Math. Methods Med., 2011, 2011, 527610

34. von M. Sahini and M. Sahimi, Applications Of Percolation Theory, CRC Press, 2003

35. D. Stauffer and A. Aharony, Introduction to percolation theory, 2nd, CRC Press, 1994

36. S. Davis, P. Trapman, H. Leirs, M. Begon, and J. A. P. Heesterbeek, The abundance threshold for plague as a critical percolation phenomenon, Nature, 2008, 454, 634–637

37. R. Parshani, S. Carmi, and S. Havlin, Epidemic Threshold for the Susceptible-Infectious-Susceptible Model on Random Networks, Phys. Rev. Lett., 2010, 104, 258701

38. Q. Li, X. Guan, P. Wu, et al. Early transmission dynamics in Wuhan, China, of novel coronavirus-infected pneumonia, N. Engl. J. Med., 2020, DOI:10.1056/NEJMoa2001316

39. M. Read, J. RE Bridgen, D. AT Cummings, A. Ho, C. P. Jewell, Novel coronavirus 2019-nCoV: early estimation of epidemiological parameters and epidemic predictions, medRxiv, 2020, DOI: https://doi.org/10.1101/2020.01.23.20018549

40. J. T. Wu, K. Leung, and G. M. Leung, Nowcasting and forecasting the potential domestic and international spread of the 2019-nCoV outbreak originating in Wuhan, China: a modeling study, The Lancet, 2020, DOI:https://doi.org/10.1016/S0140-6736(20)30260-9

41. T. Hao, Y. Xu, and Ting Hao, Exploring the Inflation and Gravity of the Universe with Eyring’s Rate Process Theory and Free Volume Concept, Physics Essays, 2018, 31(2), 177–187

